# Reduced nighttime smartphone use among cohabiting partners: a longitudinal study under the lens of social control of health behaviors theory

**DOI:** 10.64898/2026.06.09.26355243

**Authors:** Tove A. Klasson, Naja Hulvej Rod, Adrian G. Zucco

## Abstract

**Objective:** We examined the link between cohabitation with a partner and nighttime smartphone use through the social control of health behavior theory.

**Background:** Nighttime smartphone use is a behavioral risk factor for sleep problems. While previous research has predominantly focused on individual-level risks of sleep disturbances, the role of social context remains underexplored. Theoretical frameworks, specifically the Social Control of Health Behavior, suggest that social relationships regulate health-related behaviors; however, it is unclear how far this regulation extends to modern digital behaviors among couples.

**Method:** We analyzed survey data from three waves of the SmartSleep Study (2018, 2020, and 2023; total N = 25,028), including a longitudinal follow-up subset (N = 1,003). We tested multivariate associations between living with a partner, changes in cohabitation status and frequent nighttime smartphone use by fitting generalized linear mixed-effects models. Additionally, we mapped the complex interplay between indicators of social integration, social support, smartphone use, and sleep quality using hierarchical clustering of non-linear correlations.

**Results:** Cohabiting participants had lower odds of frequent nighttime smartphone use compared to those living alone (OR = 0.66; 95% CI: 0.61, 0.72). This lower risk was driven primarily by cohabitation with a partner (OR = 0.49; 95% CI: 0.36, 0.66). Longitudinal analysis supported these findings, showing that sustained cohabitation was associated with less frequent nighttime use (OR = 0.56; 95% CI: 0.38, 0.82). Clustering analysis revealed that indicators of social integration and support clustered with favorable sleep quality.

**Conclusion:** Our findings suggest that the health-protective effects of cohabitation with a partner extend to digital behaviors. Consistent with social control of health behavior theory, the presence of a partner appears to reduce frequent nighttime smartphone use, highlighting the critical importance of considering social context when addressing digital health hygiene and promoting sleep.

## Introduction

Sleep is critical for human health and well-being [1], and ignoring sleep problems could lead to serious public health issues [2]. Globally, approximately 16 percent of adults suffer from insomnia [3], with prevalence reaching up to 25 percent in some countries [4]. While sleep problems are complex and multifactorial, the widespread adoption of smartphones has emerged as an important environmental and behavioral risk factor [5]. Unlike many traditional sleep disruptors, smartphone use is a modifiable behavior, making it a particularly relevant target for public health intervention.

Smartphones are a relatively new addition to people’s lives, fundamentally altering daily behaviors, including how we communicate and entertain ourselves. One particular type of behavior that has drawn attention is nighttime smartphone use [6]. This is because the period surrounding sleep is biologically and behaviorally sensitive: disruptions by smartphone use during this window can delay sleep onset, reduce sleep duration, and compromise sleep quality [6], [7], [8], [9], [10], ultimately impacting overall health [11]. Nighttime smartphone use is highly prevalent, particularly among adolescents and young adults [12], [13], which amplifies its public health relevance [14]. Two main mechanisms are typically proposed to explain why this behavior translates into negative health outcomes: physiological disruption due to exposure to bright light from the screen [15] and cognitive arousal due to the content consumed [16], [17]. However, there is a lack of consensus regarding the exact physiological magnitude of screen light’s effect on sleep at the individual level [18] and understanding the underlying psychological mechanisms requires a broader look beyond the individual [19]. This points to an important question: to what extent do the social contexts in which people use their smartphones at night shape this behavior and could these contexts offer a lever for prevention?

One such context is the shared sleeping environment. Research consistently shows that social relationships are positively associated with health in general [20] and with sleep quality [21]. This social dimension extends to digital behaviors: evidence suggests that certain social contexts can mitigate harmful nighttime smartphone use [22]. Among cohabiting couples, for instance, disturbance from a partner’s smartphone use in bed is common and seen as an accepted part of modern life. To adapt, couples often change their nighttime smartphone behaviors and adopt strategies to avoid disturbing their partner’s sleep [10]. The theory of Social Control of Health Behaviors by Umberson [23] provides a theoretical framework for understanding this dynamic by explaining how people in social relationships positively influence each other’s health behaviors. Through this mechanism, we propose that cohabitation with a partner may function as a protective social context that reduces the frequency of nighttime smartphone use.

In this study, we examine the link between cohabitation with a partner and nighttime smartphone use, building upon the theory of Social Control of Health Behavior [23]. We utilized large-scale survey data from three waves of the SmartSleep Study in 2018, 2020, and 2023 [24], comprising a total cross-sectional sample of 25,135 individuals and a longitudinal follow-up subset of 1,003 participants. First, we aimed to determine the cross-sectional and longitudinal associations between living with a partner, changes in cohabitation status over time and frequent nighttime smartphone use. Second, we sought to map the broader, complex interplay between indicators of social integration, social support, smartphone use, and sleep quality. Consistent with the Social Control of Health Behavior framework, we hypothesized that cohabiting with a partner would exert a health-protective effect by reducing the frequency of nighttime smartphone use, thereby highlighting the critical role of social context in promoting digital health hygiene and improving overall sleep quality.

## Methods

### Study population

The study used three waves of the SmartSleep Study [24], specifically the Citizen Science Sample (2018) as a baseline sample and two waves of follow-up (2020 and 2023) for a subset of the cohort, with specific questions on social relationships.

**Figure 1.**
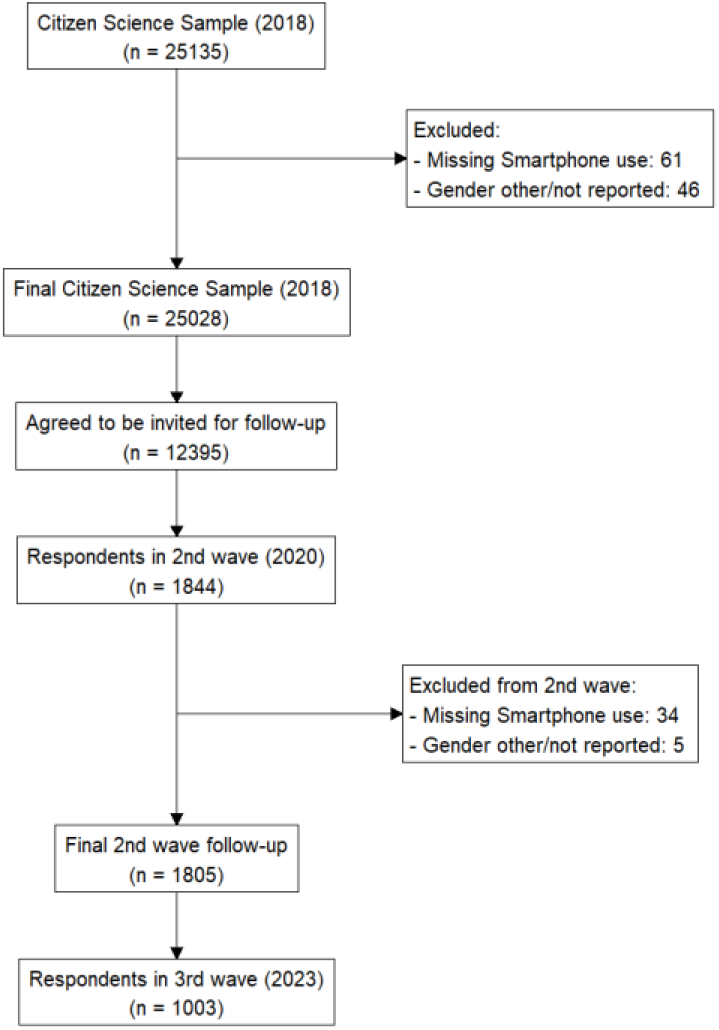
Flow chart of the Citizen Science sample and follow-up waves.

The Citizen Science sample was collected during one week in November 2018 in collaboration with the Danish national radio. A total of 25,135 Danish citizens answered a survey concerning their sleep health and smartphone behavior. See detailed descriptions of the sampling process in the cohort profile [24].

For our analysis, we excluded those without reported smartphone use and those who identified as a gender that was neither “male” nor “female”. The exclusion due to gender was due to privacy issues with low sample sizes compromising the privacy of the participants. Our primary analysis was based on the resulting sample of 25,028 individuals. Among the 12,395 who agreed to be invited for follow-up surveys, 1,805 responded and passed the inclusion criteria contributing to the second wave of follow-up from July 2020. The third wave of follow-up data aimed at gathering more detailed information about the role of social and romantic relationships in sleep and smartphone use. Data were collected in November 2023 and from those in the second wave, 1,003 were included in the sample. The follow up samples with detailed questions regarding co-habitation were used for assessing the effect of changes in co-habitation status and non-linear correlations with frequent nighttime smartphone use.

All data collection in the SmartSleep Study has received approval under the University of Copenhagen’s umbrella authorization from the Danish Data Protection Agency (datatilsynet.dk). This ensures compliance with both national and EU data security regulations for the citizen science sample (no. 514-0237/18-300) and the subsequent waves of follow-up (no. 514-0288/19-3000). Participation in all aspects of the study was voluntary.

### Nighttime smartphone usage

Frequent nighttime smartphone usage was our main outcome based on the answers to a survey question of nighttime smartphone use: how often the participant used their smartphone after sleep onset in the last three months [24]. The answering options were coded as Yes or No based on the following answers: 1. “Every night or almost every night” (Yes), 2. “Several nights a week” (Yes); 3. “Several nights a month or less” (No); and “Never” (No).

### Co-habitation status

In the Citizen Science sample, participants were asked if they live alone. Negative responses to this question indicated that the participant lived with others but lacked information about the type of social relation with the other co-habitants. In the follow-up survey waves, specific questions were asked about whom the participants live with, including romantic partners. We consider these variables the most indicative measures available in our data of whether or not an individual is in a social context at night.

### Social control of nighttime smartphone use

Social control of health behaviors is itself a complex process influenced by both the intentions and the perceptions of the individuals involved, as well as the strategies employed, which can yield positive, neutral, or negative effects [25]. While negative effects are possible, previous studies point mainly to positive effects [20], [25], [26]. The theory served as the guiding framework for our investigation and analysis of smartphone usage and relationship status. Two key concepts from Social Control of Health Behavior theory made up our framework: social integration and social support. Social integration refers to the level of involvement with social relationships (both informal and formal) [27]. Social support refers to the emotional qualities and functions of a relationship [25], [27].

Based on these two key concepts, we selected variables that reflected concrete aspects of social integration: Relationship status, Living situation (with whom the participants live), Co-sleeping situation (with whom the participants co-slept), Number of close friends, and Frequency of contact with friends. The variables used to reflect social support pointed to indirect and emotional aspects of social relationships: Feeling like you have someone to talk to, Relationship satisfaction, and the short 3-item UCLA Loneliness Scale version [28]. Based on our theoretical framework, we expected all of these aspects to act as protective factors against nighttime smartphone use, but through the two different mechanisms of integration and support, thus potentially leading to different outcomes [25], [26].

### Baseline covariates

In the first phase of the analysis, the following control variables were included: age, gender (male/female), education, mobile dependency, perceived stress and survey year. These variables were included because of their established associations with health behaviors in general, and sleep health specifically [4], [9], [29], [30], [31], [32], [33], [34], [35]. The age variable was coded into categories of ten years. The gender variable was encoded into two categories, “Female” and “Male”.

The highest completed education determined the education variable. To ensure a sufficient sample size for each category, the original survey response options were grouped into three primary categories: 1) “Basic Education” (comprising primary school), 2) “Secondary Education” (merging technical/vocational education and upper secondary education), and 3) “Higher Education” (merging short, medium, and long cycle higher education). Additionally, “Current student” was identified using a separate survey question regarding current occupation, and “Other” was maintained as a separate category.

The mobile dependency variable was based on the seven statements making up the Problematic Mobile Phone Use Questionnaire (PMPU) [24], [36] and stress based on the 10-item perceived stress scale [37]. These scales were included as indicators of mental health [38].

### Statistical analysis

First, we computed descriptive summary statistics for each of the samples used in our study (Table 1). Then, to investigate the association between co-habitation status and frequent nighttime smartphone use, we utilized generalized linear mixed-effects models (GLMMs) specifying a binomial response distribution with a logit link (mixed-effects logistic regression). A random intercept for each participant was included to account for the non-independence of repeated measures across survey waves.

**Table 1.**
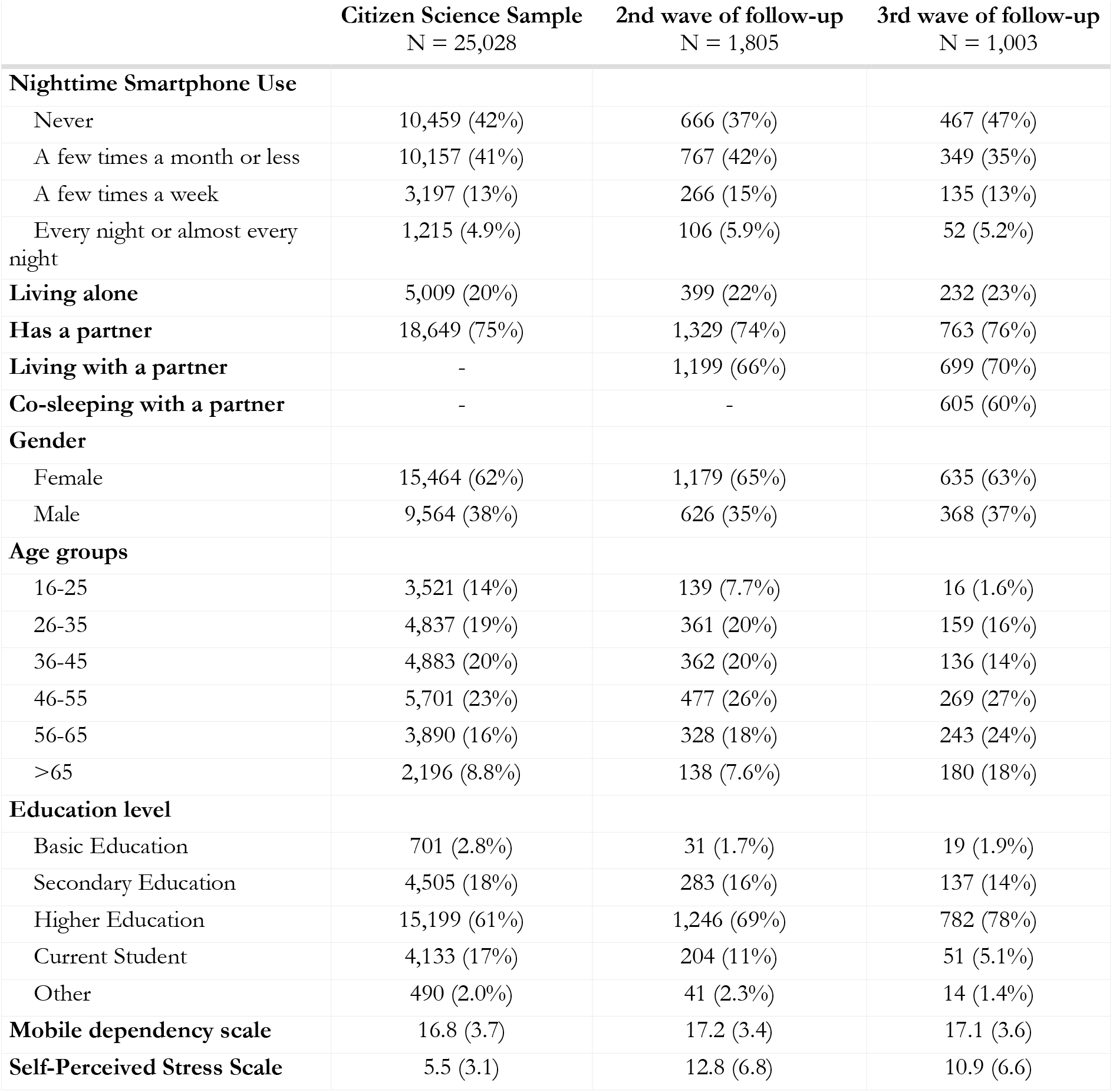
Descriptive statistics of the Citizen Science (2018), second (2020) and third wave (2023) of follow-up samples from the SmartSleep study. n (%); Mean (SD)

The primary analysis was structured around two main models. In the first model, we examined the association between living alone and frequent nighttime smartphone use across all available survey rounds. In the second model, we focused specifically on the impact of living with a partner, utilizing data restricted to the second and third follow-up waves from 2020 and 2023. Both models were adjusted for the previously described baseline covariates: age group, gender, education level, mobile dependency, perceived stress, and the specific survey year. Results from these models were reported as Odds Ratios (OR) with corresponding 95% Confidence Intervals (CI).

Furthermore, we conducted a longitudinal transition analysis to evaluate how changes in living status over time impacted nighttime smartphone usage. Using data from 2020 and 2023 waves, we categorized participants into four distinct transition groups based on their relationship status at both time points. These categories comprised individuals who had no partner at either time point, those who lived with a partner consistently, those who moved in with a partner between the two waves, and those who stopped living with a partner. A similar mixed-effects logistic regression model was then applied to assess the association between these relationship transition statuses and frequent nighttime smartphone usage at the end of the follow-up period. This transition model was adjusted for age, education, gender, mobile dependency, and perceived stress.

### Non-linear correlation structure through hierarchical clustering

To explore potential complex interactions among variables related to social control of health behavior, smartphone use, and sleep, we computed non-linear pairwise correlations using data from the third wave of follow-up. All variables reflecting social relationships were explored together with sleep quality and all available variables concerning smartphone behavior. In total, 30 variables were included in the analysis (Supplementary Table 1). We computed pairwise Kendall correlations between all variables, which we appropriately encoded based on their data type (categorical, ordinal, or continuous) [39]. We then transformed the resulting correlation matrices into distance matrices, defined as 1−*ρ*. These distance matrices were subsequently processed using agglomerative hierarchical clustering based on the unweighted pair group method with arithmetic mean (UPGMA). We depicted the outcome of this hierarchical clustering approach as a dendrogram, a tree-like diagram that visually represents the hierarchical relationships between variables. In a dendrogram, branches indicate the sequence in which variables were merged, and the height of each merge point (node) signifies the dissimilarity at which the groups of variables were combined. This approach allowed us to comprehensively explore the complex structural relationships within the variables of interest. This meant we could identify variables that are relatively grouped together due to strong non-linear correlations, thereby revealing factors closely correlated with both poor and good sleep quality while considering various dimensions of social control. The analysis was performed using R 4.4.1 with the tidyverse, corrplot, and dendextend libraries.

## Results

### Baseline characteristics

We found similar distributions of nighttime smartphone use, mobile dependency, gender, and co-habitation status among the citizen science and follow-up samples (Table 1). In the third wave of follow-up, the participants were slightly older with more of them having completed higher education. Overall, the individuals were most likely to be middle-aged, female, have a higher education, at least occasionally use their phone at night, live with a partner, and have similar levels of mobile dependency in all three samples. Perceived stress was higher in the follow-up samples collected in 2020 and 2023, which might be explained by the samples being collected during the start and end of the COVID-19 pandemic.

### Lower nighttime smartphone use among people living with a partner

Living with someone is associated with lower odds of frequent nighttime smartphone usage (OR: 0.66; 95% CI: 0.61, 0.72) than living alone. When specifically examining the social relation with the co-habitants in the follow-up waves, living with a partner was associated with even lower odds of frequent nighttime smartphone use (OR: 0.49; 95% CI: 0.36, 0.66) compared to living alone, while living with others showed no clear association (Figure 2).

**Figure 2.**
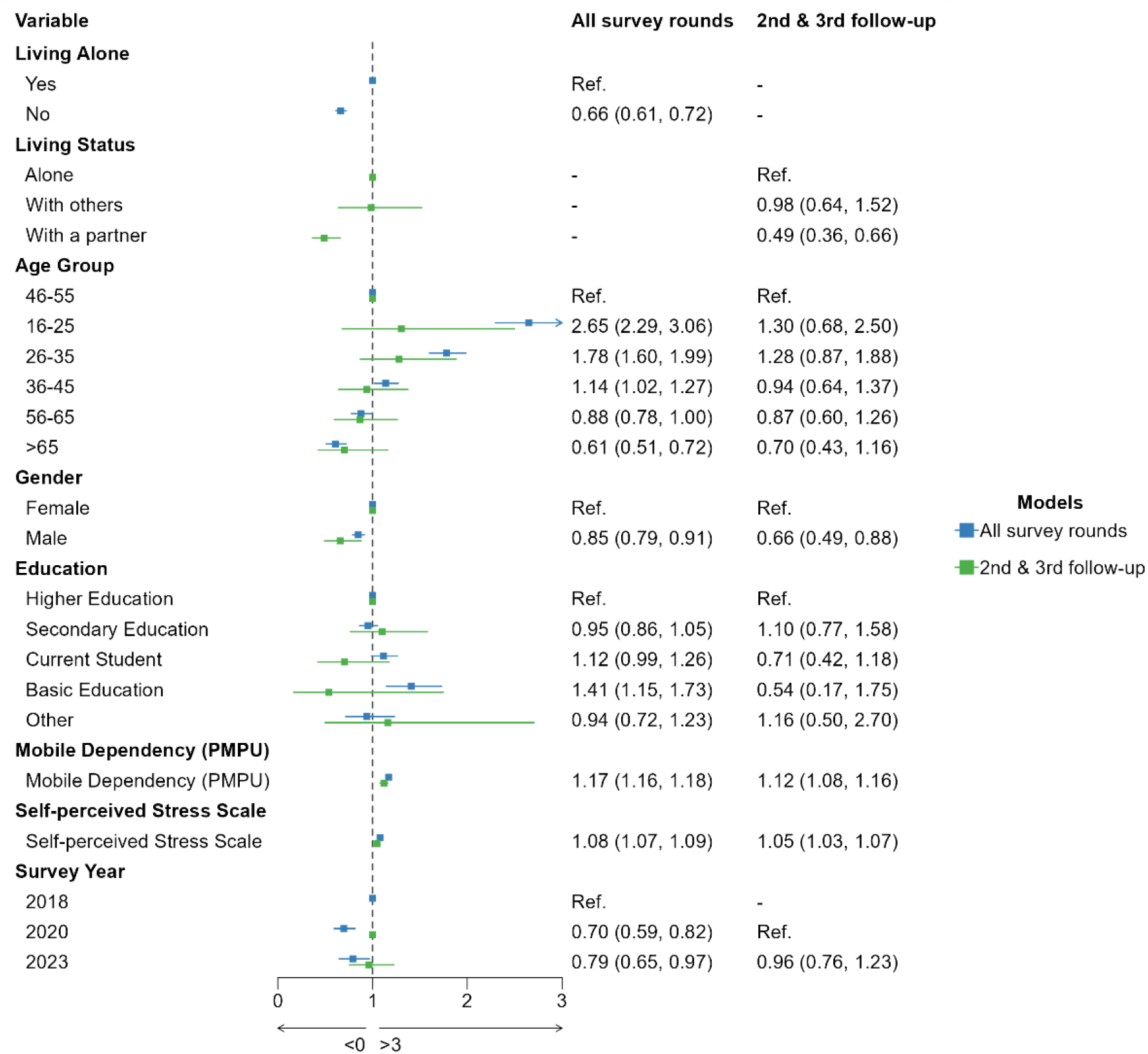
Multivariable forest plot of factors associated with frequent nighttime smartphone use. Odds Ratios (OR) and 95% Confidence Intervals (CI) were derived from generalized linear mixed-effects models adjusting for age, gender, education, mobile dependency (PMPU), perceived stress, and survey year. The blue markers represent the analysis of all survey rounds, while the green markers represent the analysis focused on the second and third follow-up rounds. Random intercepts were included for participant ID to account for repeated measures.

We found lower odds of frequent nighttime smartphone use in participants over 65 years old (OR: 0.61; 95% CI: 0.51, 0.72) and higher odds among those aged 16–25 (OR: 2.65; 95% CI: 2.29, 3.06). We found no association between education and frequent nighttime smartphone use, except for those with basic education, who had higher odds of nighttime smartphone use (OR: 1.41; 95% CI: 1.15, 1.73) compared to those with higher education. Respondents identifying as female were more likely to have frequent nighttime smartphone use, and both mobile dependency (OR: 1.17; 95% CI: 1.16, 1.18) and self-perceived stress (OR: 1.08; 95% CI: 1.07, 1.09) were associated with higher risks of frequent nighttime smartphone use. Finally, the odds of frequent nighttime smartphone use were lower in the 2020 and 2023 survey rounds compared to the 2018 baseline.

### Longitudinal shifts in co-habitation status show protective effects of living with a partner on nighttime smartphone use

To understand the dynamics of living arrangements over time, we tracked participant living status across all three survey waves (2018, 2020, and 2023). The longitudinal flow (Figure 3) illustrates a high degree of stability in living arrangements, though transitions occurred between surveys, particularly regarding moving in with or away from partners. Despite lacking details on the social relationship with the co-cohabitants in 2018 we could expect that most participants not living alone would be living with a partner.

**Figure 3.**
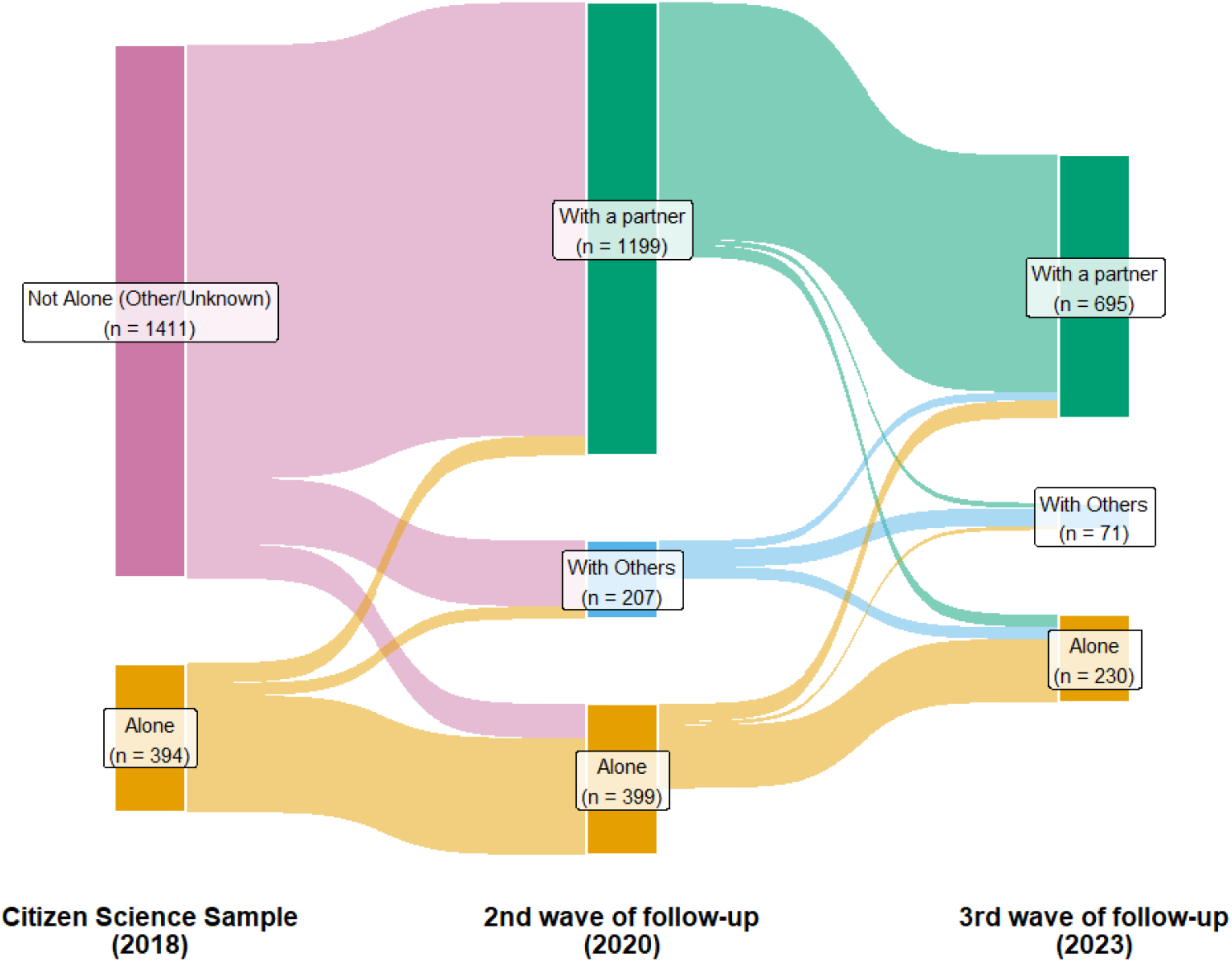
Longitudinal transitions in co-habitation status across three study waves (2018–2023). Sankey diagram visualizing the flow of participants between different living arrangements: “Alone”, “Not Alone”, “With a partner” and “With Others”. The social relationship with co-habitants was unknown in 2018. The node labels indicate the total number of participants in each category at each time point. The 2018 wave represents the initial Citizen Science sample, followed by the second (2020) and third (2023) waves of follow-up for a maximum of 1805 participants.

We further analyzed how these specific transitions influenced nighttime smartphone behaviors between 2020 and 2023, for which information on co-habitation with a partner was available. Living arrangements with a partner were associated with lower odds of frequent nighttime smartphone use (Table 2). Specifically, participants who consistently lived with a partner throughout the follow-up period had lower odds of frequent nighttime use compared to those who remained without a partner (OR = 0.56; 95% CI: 0.38, 0.82).

**Table 2.** Association between transitions in co-habitation status and frequent nighttime smartphone use. Odds Ratios (OR) were adjusted for age, education, gender, PMPU scale, and stress scale. CI = Confidence Interval.

| Status | N | Events | OR | 95% CI |
| --- | --- | --- | --- | --- |
| No partner | 250 | 68 | — | — |
| Living with a partner | 598 | 91 | 0.56 | 0.38, 0.82 |
| Moved in with a partner | 65 | 12 | 0.48 | 0.23, 1.00 |
| Stopped living with a partner | 45 | 6 | 0.42 | 0.16, 1.08 |

While participants who moved in with a partner (OR = 0.48; 95% CI: 0.23, 1.00) or stopped living with a partner (OR = 0.42; 95% CI: 0.16, 1.08) during this period also showed a trend toward lower nighttime smartphone use compared to those who remained alone, these results were not robust likely due to the smaller sample sizes in these transition groups.

### The complex relationships between social integration, social support, smartphone use, and sleep quality

Guided by the theory of Social Control of Health Behavior, we selected indicators of social integration, social support, smartphone-related behaviors, and sleep health available in the last follow-up sample from 2023. These concepts were represented by 30 variables from 18 survey questions (Supplementary Table 1).

The resulting dendrogram at its highest level of hierarchy reveals a separation between behavioral and psychosocial factors correlated with positive or negative sleep outcomes (Figure 4). We found a strong grouping between poor sleep quality, loneliness, and problematic smartphone behaviors. Specifically, indicators of poor sleep quality were located closely to a sub-cluster of variables representing perceived loneliness, social media use, mobile dependency, nighttime smartphone use, and feeling disturbed by the phone. This grouping suggests that feelings of lacking social support are intertwined with a high reliance on smartphones, particularly during nighttime, which collectively correlate with poor sleep health.

**Figure 4.**
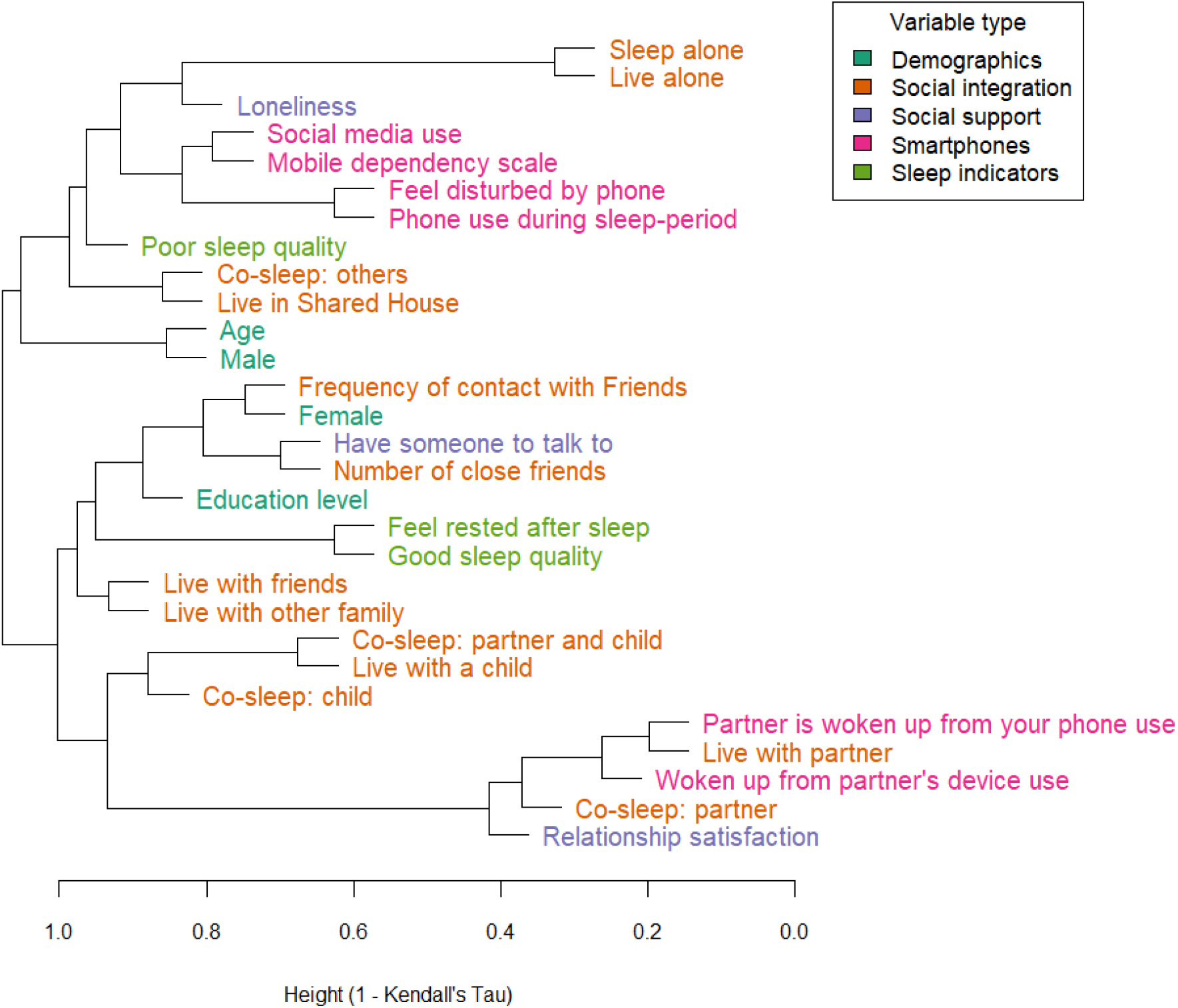
The complex relationships among indicators of social integration, social support, sleep, and smartphone use. Dendrograms of hierarchical clustering based on distance matrices defined as 1 – Kendall correlations (non-linear pairwise correlations) among indicators in the 3rd wave of follow-up.

In contrast, positive sleep outcomes clustered alongside robust indicators of social integration and support. Variables such as “Good sleep quality” and “Feel rested after sleep” form a distinct, tightly correlated pairing that is situated within a broader branch containing “Number of close friends,” “Have someone to talk to,” and “Frequency of contact with friends.” This structural divergence in the dendrogram represents a dichotomy within the data: robust, supportive social networks align with restorative sleep, whereas loneliness aligns with maladaptive digital habits and poor sleep outcomes.

In these two clusters a gender difference also becomes evident: men are more likely to live alone, sleep alone, have more nighttime smartphone usage and poor sleep quality, while women are more likely to live with friends or family, have someone to talk to and have good sleep quality. Overall, the hierarchical clustering structure provides support for our hypothesis, illustrating the important links between an individual’s social relationships, their smartphone usage patterns, and, ultimately, their sleep health.

## Discussion

In this study, we investigated the complex associations between social relationships, co-habitation status, and nighttime smartphone use across three waves of the SmartSleep study. Guided by the theory of Social Control of Health Behavior, we found that individuals living with others consistently exhibited lower odds of frequent nighttime smartphone use compared to those living alone. Furthermore, longitudinal data from the second and third survey waves revealed that living with a romantic partner was associated with the lowest nighttime device use. These results support the theory and suggest that the type and quality of a social tie are crucial; emotionally close relationships appear to foster a sense of shared responsibility that discourages maladaptive health behaviors that affect sleep hygiene.

The protective nature of these partnerships is further highlighted by our longitudinal transition analysis. By tracking changes in co-habitation status between 2020 and 2023, we found that maintaining a stable living arrangement with a partner yielded the lowest levels of frequent nighttime smartphone use compared to those who consistently lived without a partner. Interestingly, individuals who experienced relationship transitions during this period, whether moving in with a partner or stopping living with one, also indicated lower nighttime device use relative to the consistently single group. Although these transition effects were based on smaller sample sizes than our primary analysis. For instance, the lower odds among those who recently stopped living with a partner may suggest a potential “carryover” effect, where positive sleep hygiene habits established under the social control of a past relationship persist even after the co-habitation ends. These temporal dynamics highlight that stable, long-term emotional integration may help reduce frequent nighttime smartphone use.

To our knowledge, this is the first study to explicitly apply social control of health behavior theory to digital health behaviors, specifically assessing how cohabitation status impacts nighttime smartphone use. The critical importance of behavior in determining sleep health is widely recognized in clinical practice. Cognitive-behavioral therapy for insomnia (CBT-I) is recommended as the first-line treatment for adults of any age, because modifying maladaptive habits is so essential to restorative sleep [40]. By applying a theoretical framework from sociology, our findings suggest that living with others could act as a natural, interpersonal mechanism for behavioral regulation. Our regression analyses utilizing a large citizen science sample empirically confirm and extend previous research regarding the protective effects of romantic relationships on both general sleep health and smartphone use behaviors [10], [21], [41], [42]. Specifically, our findings point to how the regulatory role of social control within intimate partnerships [21], [22], [26] effectively reinforces positive, CBT-I-aligned sleep habits, such as curtailing nighttime device use.

To deeper explore the underlying patterns behind these behaviors, we utilized non-linear pairwise correlations and hierarchical clustering on a wide array of social, behavioral, and sleep variables. Within our theoretical framework, we treated measures of social integration and social support as indicators for active social control. While the regression models confirmed that cohabitation generally is associated with less device use, the clustering analysis revealed important details. We found that indicators of social integration, such as contact and the number of close friends, and living with friends, family, or a partner, clustered tightly with good sleep quality. Conversely, living in a “shared house” (representing physical proximity to others without necessarily implying close social support) clustered with nighttime smartphone usage, poor sleep quality, and loneliness.

These divergent clusters highlight the immense complexity of social dynamics. Our findings suggest that sharing a living space with individuals who provide genuine social support is protective, whereas merely co-residing with other adults is not sufficient to curb negative health behaviors. This provides a critical update to earlier literature, which posited that mere physical proximity to other adults is inherently beneficial for health [43]. By separating physical proximity from social integration and support, our analysis suggests that the protective mechanisms against nighttime smartphone use rely on the shared emotional responsibility found in close relationships, rather than simply having another person in the vicinity. Overall, this dynamic offers a modern, digital-age explanation for why couples generally exhibit better sleep health than their single counterparts [21], [22], [41], [42], while also reinforcing the well-documented association between loneliness, poor sleep, and compensatory screen time [44], [45], [46].

### Strengths and limitations

We employed integrated methods from data science and epidemiology within a relevant theoretical framework from sociology to thoroughly assess the association between relationship status and nighttime smartphone use. Utilizing panel data from three waves of the SmartSleep study allowed us to track these dynamics over time and assess changes in cohabitation status. Furthermore, deploying hierarchical clustering without imposing strong modeling assumptions enabled us to uncover complex, non-linear structures among indicators of social support, social integration, sleep quality, and digital behaviours.

However, several limitations must be acknowledged. First, while self-reported data allow for nuanced insights into participants’ experiences, it is subject to recall and reporting biases. Second, the study sample is not entirely representative of the broader Danish population; females, younger individuals, and those with higher educational attainment were overrepresented. Third, participants who selected “other” for their gender identity had to be excluded from the regression and clustering analyses due to low numbers. Given that gender emerged as an important factor in nighttime smartphone use, future sleep research must prioritize multidimensional approaches to gender to better capture the biopsychosocial factors influencing health behaviors [47]. Fourth, our indications for social integration and social support do not capture the full mechanisms of social control; they indicate the potential presence of social control. Future studies in smartphone use should include more in-depth information on why and how people make the behavior choices they make.

Additionally, sleep quality and mental health are intrinsically linked, with sleep disturbances often serving as both a predictor and diagnostic criterion for conditions like depression [4], [38], [48]. While our primary models adjusted for perceived stress to account for this overlap, the potential for residual confounding from unmeasured psychiatric variables remains. Finally, while our non-linear correlations effectively mapped the relational structures between variables, these associations cannot be interpreted as strictly causal, nor have they been adjusted by relevant covariates. Future longitudinal research utilizing larger datasets is required to further stratify feelings of loneliness across distinct relationship statuses and directly test the causal pathways of social control on digital habits.

## Conclusion

Our findings indicate that social relationships, specifically those characterized by strong social support and emotional integration, play a central role in regulating nighttime smartphone use. Although device use is frequently conceptualized as an isolated, individual behavior, our results emphasize that important social regulatory mechanisms have been largely overlooked. Recognizing the influence of the social and domestic context on digital behaviors has important implications for sleep outcomes. We propose that public health interventions aimed at improving sleep hygiene should shift away from solely individualistic approaches and instead incorporate strategies centered on social contexts and fostering relations, thereby mitigating the underlying drivers of excessive nighttime screen time.

## Author statement

Conceptualization: T.A.K, A.G.Z. and N.H.R. Funding acquisition: N.H.R. Methodology: T.A.K, A.G.Z., and N.H.R. Investigation: T.A.K, A.G.Z., and N.H.R. Formal analysis: T.A.K, A.G.Z. Software: T.A.K, A.G.Z. Visualization: A.G.Z. Supervision: A.G.Z. Writing—original draft: T.A.K and A.G.Z. Writing— review and editing: T.A.K, A.G.Z., and N.H.R.

We thank Christopher Jamil de Montgomery for feedback on early versions of this manuscript.

## Acknowledgments

This project was supported by funding from the Lundbeck Foundation (grant number R396-2022–352) and conducted at the Copenhagen Health Complexity Center supported by the Tryg Foundation.

## Data availability statement

The dataset contains personal identifiable data and sensitive information. We are therefore not allowed to make them publicly available according to the Danish Protection Agency (Danish data protection legislation datatilsynet.dk) and Danish law. Inquiries for secure access under conditions stipulated by the Danish Data Protection Agency should be directed at the principal investigator of the SmartSleep Study Professor Naja Hulvej Rod

## Declaration of generative AI and AI-assisted technologies in the manuscript preparation process

During the preparation of this work, A.G.Z. used a large language model (Gemini Pro 3.1) to assist with proofreading and to suggest improvements to the manuscript and software. After using this tool/service, all authors reviewed and edited the content as needed and take full responsibility for the content of the published article.

## Supplementary

**Supplementary Table 1.** Variables included in the clustering analysis and coding of the responses.

| Variable | Survey questions | Response type | Response options | Ordinal coding |
| --- | --- | --- | --- | --- |
| **Age** | How old are you? | Open field with restrictions | >15 | As original |
| **Gender** | What is your gender? |  | 1. Male<br>2. Female<br>3. Other | Male = 0, Female = 1, Other = 2 |
| **Live alone** | Do you live alone? |  | 1. Yes<br>2. No | Yes = 1, No = 0 |
| **Live with...** | Who are you living with? | Multiple responses possible | 1. Spouse/girlfriend/boyfriend<br>2. child/children<br>3. other family<br>4. friend(s)/roommate(s)<br>5. shared house, college, continuation school, folk high school<br>6. Alone | Separate dummy variables:<br>Yes = 1, No = 0 |
| **Has a partner** | Do you have a romantic partner? |  | 1. Yes<br>2. No | Yes = 1, No = 0 |
| **Co-sleep with...** | Who are you co-sleeping with? | Multiple responses possible | 1. Alone<br>2. Partner<br>3. Child<br>4. Partner and child<br>5. Others | Separate dummy variables:<br>Yes = 1, No = 0 |
| **Number of close friends** | How many close friends do you have? |  | 1. 10 or more<br>2. 6-9<br>3. 3-5<br>4. 2<br>5. 1<br>6. "Don't know"<br>7. "No close friends" | "No close friends" = -1,<br>"Don't know" = 0,<br>1 = 1,<br>2 = 2,<br>3-5 = 3,<br>6-9 = 4,<br>10 or more = 5 |
| **Frequency of contact with friends** | How often are you in contact with friends and family that you do not live with?<br>In contact with is defined by being together, talking in the phone with each other or texting each other etc. |  | 1. Every day or almost every day<br>2. Every week<br>3. Every month<br>4. Less than every month<br>5. Never | "Never" = 0,<br>"Less than every month" = 1,<br>"Every month" = 2,<br>"Every week" = 3,<br>"Every day or almost every day" = 4 |
| **Loneliness** | UCLA short 3-item loneliness scale [28]. | Scale | 3-9 | As original |
| **Have someone to talk to** | Do you have anyone to talk to if you have any problems or need support? |  | 1. Yes, always<br>2. Yes, most of the time<br>3. Yes, sometimes<br>4. No, never or almost never | “No, never or almost never” = 0,<br>“Sometimes” = 1,<br>“Most of the time” = 2,<br>“Always” = 3 |
| **Relationship satisfaction** | Overall, how satisfied are you with your relationship? |  | 1. Very dissatisfied,<br>2. Fairly dissatisfied,<br>3. Neither satisfied or dissatisfied,<br>4. Fairly satisfied,<br>5. Very satisfied | “Very dissatisfied” = 0,<br>“Fairly dissatisfied” = 1,<br>“Neither satisfied or dissatisfied” = 2,<br>“Fairly satisfied” = 3,<br>“Very satisfied” = 4 |
| **Sleep quality** | Karolinska Sleep Questionnaire [49] | Scale | 1-5 | As original,<br>reversed for good sleep quality |
| **Mobile dependency** | the Problematic Mobile Phone Use Questionnaire [24], [36]. | Scale | 7-28 | As original |
| **Social Media use** | How often do you use social media? |  | 1. Never<br>2. Weekly or less<br>3. Once a day<br>4. Several times a day<br>5. Every second hour<br>6. Every hour | “Never” = -1,<br>“Weekly or less” = 0,<br>“Once a day” = 1,<br>“Several times a day” = 2,<br>“Every second hour” = 3,<br>“Every hour” = 4 |
| **Disturbed by phone** | In general, do you feel disturbed by your mobile phone after you went to sleep? |  | 1. Never<br>2. Rarely<br>3. Sometimes<br>4. Often<br>5. Very often | “Never” = 0,<br>“Rarely” = 1,<br>“Sometimes” = 2,<br>“Often” = 3,<br>“Very often” = 4 |
| **Phone use during sleep period** | Think of the last three months. How often did you check your mobile phone after you went to sleep?<br>”To check” refers to both short and long activation of your mobile phone e.g. turn on the screen or use of apps. |  | 1. Never<br>2. Few times a month or less<br>3. Few times a week<br>4. Every night or almost every night | “Never” = 0,<br>“A few times a month or less” = 1,<br>“A few times a week” = 2,<br>“Every night or almost every night” = 3 |
| **Woken up from partner's device use** | Are you awakened by your partner who uses his/her mobile phone or other digital devices in bed? |  | 1. Never<br>2. Sometimes<br>3. Often<br>4. My partner does not use digital devices in the bed<br>5. No partner | "No partner" = -1.<br>"My partner does not use digital devices in the bed" = 0,<br>"Never" = 0,<br>"Sometimes" = 2,<br>"Often" = 4 |
| **Partner is woken up from your phone use** | Does your partner complain about being woken up by your mobile phone use in bed? |  | 1. Never<br>2. Sometimes<br>3. Often<br>4. I don't use digital devices in bed<br>5. No partner | "No partner" = -1.<br>"I don't use digital devices in bed" = 0,<br>"Never" = 0,<br>"Sometimes" = 2,<br>"Often" = 4 |
| **Education level** | Current highest completed education |  | 1. Primary school<br>2. Upper secondary education<br>3. Technical/Vocational education<br>4. Short cycle higher education<br>5. Medium cycle higher education<br>6. Long cycle higher education<br>7. Other | "Primary School" = 1<br>"Upper Secondary Education" = 2,<br>"Technical Vocational Education" = 2,<br>"Short Cycle Higher Education" = 3,<br>"Medium Cycle Higher Education" = 4,<br>"Long Cycle Higher Education" = 5<br>"Other" = 0 |
| **Feel rested after sleep** | Do you think you get enough sleep to feel rested? |  | 1. No<br>2. Yes, but not often enough<br>3. Yes, usually | "No" = 0,<br>"Yes, but not often enough" = 1,<br>"Yes, usually" = 2 |

**Supplementary Table 2.** Logistic regression analysis of factors associated with frequent nighttime smartphone use among cohabiting couples. Results are presented as Odds Ratios (OR) with 95% Confidence Intervals (CI). The model examines the primary association with co-sleeping status, adjusted for age, education, gender, problematic mobile phone use (PMPU) scale, and stress levels among individuals in the 3^rd^ wave of follow up sample.

| Characteristic | OR | 95% CI |
| --- | --- | --- |
| Co-sleeping with a partner |  |  |
| No | — | — |
| Yes | 0.59 | 0.34, 1.04 |
| Age Group |  |  |
| 46-55 | — | — |
| 16-25 | 0.00 |  |
| 26-35 | 1.24 | 0.62, 2.44 |
| 36-45 | 0.56 | 0.26, 1.18 |
| 56-65 | 0.86 | 0.46, 1.60 |
| >65 | 0.86 | 0.42, 1.72 |
| Education |  |  |
| Higher Education | — | — |
| Secondary Education | 0.79 | 0.38, 1.52 |
| Current Student | 0.95 | 0.25, 3.01 |
| Basic Education | 0.00 |  |
| Other | 3.33 | 0.61, 15.4 |
| Gender |  |  |
| Female | — | — |
| Male | 0.71 | 0.43, 1.15 |
| Mobile Dependency (PMPU) | 1.14 | 1.07, 1.22 |
| Self-perceived Stress Scale | 1.05 | 1.02, 1.09 |

## References

[1] D. J. Buysse, “Sleep Health: Can We Define It? Does It Matter?,” Sleep, vol. 37, no. 1, pp. 9–17, Jan. 2014, doi: 10.5665/sleep.3298.

[2] D. C. Lim et al., “The need to promote sleep health in public health agendas across the globe,” The Lancet Public Health, vol. 8, no. 10, pp. e820–e826, Oct. 2023, doi: 10.1016/S2468-2667(23)00182-2.

[3] A. V. Benjafield et al., “Estimation of the global prevalence and burden of insomnia: a systematic literature review-based analysis,” Sleep Medicine Reviews, vol. 82, p. 102121, Aug. 2025, doi: 10.1016/j.smrv.2025.102121.

[4] E. Aernout et al., “International study of the prevalence and factors associated with insomnia in the general population,” Sleep Medicine, vol. 82, pp. 186–192, Jun. 2021, doi: 10.1016/j.sleep.2021.03.028.

[5] S. Thomée, A. Härenstam, and M. Hagberg, “Mobile phone use and stress, sleep disturbances, and symptoms of depression among young adults - a prospective cohort study,” BMC Public Health, vol. 11, no. 1, p. 66, Jan. 2011, doi: 10.1186/1471-2458-11-66.

[6] A. S. Dissing, T. O. Andersen, L. N. Nørup, A. Clark, M. Nejsum, and N. H. Rod, “Daytime and nighttime smartphone use: A study of associations between multidimensional smartphone behaviours and sleep among 24,856 Danish adults,” Journal of Sleep Research, vol. 30, no. 6, p. e13356, 2021, doi: 10.1111/jsr.13356.

[7] T. O. Andersen et al., “Nighttime smartphone use, sleep quality, and mental health: investigating a complex relationship,” Sleep, vol. 46, no. 12, p. zsad256, Dec. 2023, doi: 10.1093/sleep/zsad256.

[8] L. AS. Brautsch, L. Lund, M. M. Andersen, P. J. Jennum, A. P. Folker, and S. Andersen, “Digital media use and sleep in late adolescence and young adulthood: A systematic review,” Sleep Medicine Reviews, vol. 68, p. 101742, Apr. 2023, doi: 10.1016/j.smrv.2022.101742.

[9] N. Liu, J. Wang, and W. Zang, “The Impact of Sleep Determination on Procrastination before Bedtime: The Role of Anxiety,” IJMHP, vol. 26, no. 5, pp. 377–387, 2024, doi: 10.32604/ijmhp.2024.047808.

[10] T. Salmela, A. Colley, and J. Häkkilä, “Together in Bed?: Couples’ Mobile Technology Use in Bed,” in Proceedings of the 2019 CHI Conference on Human Factors in Computing Systems, Glasgow Scotland Uk: ACM, May 2019, pp. 1–12. doi: 10.1145/3290605.3300732.

[11] H. J. Drews, C. Sejling, T. O. Andersen, T. V. Varga, A. K. Jensen, and N. H. Rod, “Tracked and self-reported nighttime smartphone use, general health, and healthcare utilization: results from the SmartSleep Study,” Sleep, p. zsae024, Feb. 2024, doi: 10.1093/sleep/zsae024.

[12] B. Brosnan, J. J. Haszard, K. A. Meredith-Jones, S.-R. Wickham, B. C. Galland, and R. W. Taylor, “Screen Use at Bedtime and Sleep Duration and Quality Among Youths,” JAMA Pediatrics, Sep. 2024, doi: 10.1001/jamapediatrics.2024.2914.

[13] N. H. Rod, A. S. Dissing, A. Clark, T. A. Gerds, and R. Lund, “Overnight smartphone use: A new public health challenge? A novel study design based on high-resolution smartphone data,” PLOS ONE, vol. 13, no. 10, p. e0204811, Oct. 2018, doi: 10.1371/journal.pone.0204811.

[14] A. G. Zucco, H. J. Drews, J. F. Uleman, S. Bhatt, and N. H. Rod, “Exploring nationwide patterns of sleep problems from late adolescence to adulthood using machine learning,” Science Advances, vol. 11, no. 39, p. eadw1227, Sep. 2025, doi: 10.1126/sciadv.adw1227.

[15] N. D. Dautovich et al., “A systematic review of the amount and timing of light in association with objective and subjective sleep outcomes in community-dwelling adults,” Sleep Health, vol. 5, no. 1, pp. 31–48, Feb. 2019, doi: 10.1016/j.sleh.2018.09.006.

[16] R. L. Campbell and A. J. Bridges, “Bedtime procrastination mediates the relation between anxiety and sleep problems,” Journal of Clinical Psychology, vol. 79, no. 3, pp. 803–817, 2023, doi: 10.1002/jclp.23440.

[17] A. Tandon, P. Kaur, A. Dhir, and M. Mäntymäki, “Sleepless due to social media? Investigating problematic sleep due to social media and social media sleep hygiene,” Computers in Human Behavior, vol. 113, p. 106487, Dec. 2020, doi: 10.1016/j.chb.2020.106487.

[18] M. A. Grandner, J. M. Dzierzewski, D. Gozal, J. G. Lopos, A. N. Miller, and S. Redline, “Screen use and sleep health in children, adolescents, and adults: National Sleep Foundation consensus considerations and practical suggestions,” Sleep Health: Journal of the National Sleep Foundation, vol. 11, no. 5, pp. 560–561, Oct. 2025, doi: 10.1016/j.sleh.2025.05.002.

[19] A. M. Della Vedova, L. Covolo, M. Muscatelli, Y. Loscalzo, M. Giannini, and U. Gelatti, “Psychological distress and problematic smartphone use: Two faces of the same coin? Findings from a survey on young Italian adults,” Computers in Human Behavior, vol. 132, p. 107243, Jul. 2022, doi: 10.1016/j.chb.2022.107243.

[20] J. Holt-Lunstad, T. B. Smith, and J. B. Layton, “Social Relationships and Mortality Risk: A Meta-analytic Review,” PLoS Med, vol. 7, no. 7, p. e1000316, Jul. 2010, doi: 10.1371/journal.pmed.1000316.

[21] J.-H. Chen, L. J. Waite, and D. S. Lauderdale, “Marriage, Relationship Quality, and Sleep among U.S. Older Adults,” Journal of Health and Social Behavior, vol. 56, no. 3, pp. 356–377, Sep. 2015.

[22] H. J. Drews, R. Göder, and P. Mitkidis, “Self-control or social control - what determines sleep hygiene in bed-sharing couples?,” Sleep Sci, vol. 14, no. S 02, pp. 179–184, Jun. 2021, doi: 10.5935/1984-0063.20200095.

[23] D. Umberson, “Family status and health behaviors: social control as a dimension of social integration,” J Health Soc Behav, vol. 28, no. 3, pp. 306–319, Sep. 1987.

[24] N. H. Rod et al., “Cohort profile: The SmartSleep Study, Denmark, combining evidence from survey, clinical and tracking data,” BMJ Open, vol. 13, no. 10, p. e063588, Oct. 2023, doi: 10.1136/bmjopen-2022-063588.

[25] P. A. Thoits, “Mechanisms Linking Social Ties and Support to Physical and Mental Health,” J Health Soc Behav, vol. 52, no. 2, pp. 145–161, Jun. 2011, doi: 10.1177/0022146510395592.

[26] L. Tay, K. Tan, E. Diener, and E. Gonzalez, “Social Relations, Health Behaviors, and Health Outcomes: A Survey and Synthesis,” Applied Psych Health & Well, vol. 5, no. 1, pp. 28–78, Mar. 2013, doi: 10.1111/aphw.12000.

[27] D. Umberson and J. Karas Montez, “Social Relationships and Health: A Flashpoint for Health Policy,” J Health Soc Behav, vol. 51, no. 1_suppl, pp. S54–S66, Mar. 2010, doi: 10.1177/0022146510383501.

[28] M. E. Hughes, L. J. Waite, L. C. Hawkley, and J. T. Cacioppo, “A Short Scale for Measuring Loneliness in Large Surveys: Results From Two Population-Based Studies,” Res Aging, vol. 26, no. 6, pp. 655–672, Nov. 2004, doi: 10.1177/0164027504268574.

[29] Z. Alimoradi et al., “Gender-specific estimates of sleep problems during the COVID-19 pandemic: Systematic review and meta-analysis,” Journal of Sleep Research, vol. 31, no. 1, p. e13432, Feb. 2022, doi: 10.1111/jsr.13432.

[30] B. P. Hasler and W. M. Troxel, “Couples’ Nighttime Sleep Efficiency and Concordance: Evidence for Bidirectional Associations With Daytime Relationship Functioning,” Psychosomatic Medicine, vol. 72, no. 8, pp. 794–801, Oct. 2010, doi: 10.1097/PSY.0b013e3181ecd08a.

[31] D. Kuss, L. Harkin, E. Kanjo, and J. Billieux, “Problematic Smartphone Use: Investigating Contemporary Experiences Using a Convergent Design,” IJERPH, vol. 15, no. 1, p. 142, Jan. 2018, doi: 10.3390/ijerph15010142.

[32] O. Lopez-Fernandez et al., “Self-reported dependence on mobile phones in young adults: A European cross-cultural empirical survey,” J Behav Addict, vol. 6, no. 2, pp. 168–177, Apr. 2017, doi: 10.1556/2006.6.2017.020.

[33] D. Papadopoulos and F. A. E. Sosso, “Socioeconomic status and sleep health: a narrative synthesis of 3 decades of empirical research,” Journal of Clinical Sleep Medicine, vol. 19, no. 3, pp. 605–620, Mar. 2023, doi: 10.5664/jcsm.10336.

[34] D. Umberson, “Gender, Marital Status and The Social Control of Health Behavior,” in American Sociological Association, 1992, pp. 907–917.

[35] L.-N. Zeng et al., “Gender Difference in the Prevalence of Insomnia: A Meta-Analysis of Observational Studies,” Front. Psychiatry, vol. 11, p. 577429, Nov. 2020, doi: 10.3389/fpsyt.2020.577429.

[36] J. Billieux, M. Van Der Linden, and L. Rochat, “The role of impulsivity in actual and problematic use of the mobile phone,” Applied Cognitive Psychology, vol. 22, no. 9, pp. 1195–1210, Dec. 2008, doi: 10.1002/acp.1429.

[37] S. Cohen, T. Kamarck, and R. Mermelstein, “A Global Measure of Perceived Stress,” Journal of Health and Social Behavior, vol. 24, no. 4, pp. 385–396, 1983, doi: 10.2307/2136404.

[38] J. E. Reesen et al., “A call for transdiagnostic attention to insomnia and its treatment in mental healthcare,” Journal of Sleep Research, vol. 33, no. 5, p. e14049, Oct. 2024, doi: 10.1111/jsr.14049.

[39] C. Xiao, J. Ye, R. M. Esteves, and C. Rong, “Using Spearman’s correlation coefficients for exploratory data analysis on big dataset,” Concurrency and Computation: Practice and Experience, vol. 28, no. 14, pp. 3866–3878, 2016, doi: 10.1002/cpe.3745.

[40] D. Riemann et al., “The European Insomnia Guideline: An update on the diagnosis and treatment of insomnia 2023,” Journal of Sleep Research, vol. 32, no. 6, p. e14035, 2023, doi: 10.1111/jsr.14035.

[41] H. J. Drews et al., “Bed-Sharing in Couples Is Associated With Increased and Stabilized REM Sleep and Sleep-Stage Synchronization,” Front. Psychiatry, vol. 11, p. 583, Jun. 2020, doi: 10.3389/fpsyt.2020.00583.

[42] H. J. Drews and A. Drews, “Couple Relationships Are Associated With Increased REM Sleep—A Proof-of-Concept Analysis of a Large Dataset Using Ambulatory Polysomnography,” Front. Psychiatry, vol. 12, p. 641102, May 2021, doi: 10.3389/fpsyt.2021.641102.

[43] O. Anson, “Marital Status and Women’s Health Revisited: The Importance of a Proximate Adult,” Journal of Marriage and Family, vol. 51, no. 1, pp. 185–194, 1989.

[44] S. C. Griffin, A. B. Williams, S. G. Ravyts, S. N. Mladen, and B. D. Rybarczyk, “Loneliness and sleep: A systematic review and meta-analysis,” Health Psychology Open, vol. 7, no. 1, p. 2055102920913235, Jan. 2020, doi: 10.1177/2055102920913235.

[45] T. Matthews, A. Danese, A. M. Gregory, A. Caspi, T. E. Moffitt, and L. Arseneault, “Sleeping with one eye open: loneliness and sleep quality in young adults,” Psychol. Med., vol. 47, no. 12, pp. 2177–2186, Sep. 2017, doi: 10.1017/S0033291717000629.

[46] T. Shen et al., “Exploring the association between sleep problems and loneliness in adolescents: Potential mediating effects of rumination and resilience,” Applied Psych Health & Well, vol. 17, no. 1, p. e12620, Feb. 2025, doi: 10.1111/aphw.12620.

[47] G. R. Bauer, “Sex and Gender Multidimensionality in Epidemiologic Research,” American Journal of Epidemiology, vol. 192, no. 1, pp. 122–132, Jan. 2023, doi: 10.1093/aje/kwac173.

[48] M.-M. Zhang et al., “Sleep disorders and non-sleep circadian disorders predict depression: A systematic review and meta-analysis of longitudinal studies,” Neuroscience & Biobehavioral Reviews, vol. 134, p. 104532, Mar. 2022, doi: 10.1016/j.neubiorev.2022.104532.

[49] M. Nordin, T. Åkerstedt, and S. Nordin, “Psychometric evaluation and normative data for the Karolinska Sleep Questionnaire: Evaluation of the KSQ,” Sleep and Biological Rhythms, vol. 11, no. 4, pp. 216–226, Oct. 2013, doi: 10.1111/sbr.12024.

